## Supplementary information for "SEMA6A drives GnRH neuron-dependent puberty onset by tuning median eminence vascular permeability"

**Supplementary Table 1.** GnRH neuron distribution in E14.5 embryo heads, E18.5 brains and adult MPOA.

| E14.5 heads | *Sema6a*$^{+/+}$ | | | *Sema6a*$^{-/-}$ | | | Two-tailed unpaired Student's t test |
|---|---|---|---|---|---|---|---|
| | mean | SD | n | mean | SD | n | *p* |
| NOSE | 490.5 | 111.3 | 4 | 474.8 | 37.1 | 4 | 0.7974 NS |
| CP | 331.3 | 89.9 | 4 | 307.4 | 34.5 | 4 | 0.6360 NS |
| FB | 448.5 | 38.2 | 4 | 438.8 | 98.5 | 4 | 0.8596 NS |
| TOT | 1270.3 | 135.8 | 4 | 1220.8 | 91.0 | 4 | 0.5670 NS |
| E18.5 brains | *Sema6a*$^{+/+}$ | | | *Sema6a*$^{-/-}$ | | | Two-tailed unpaired Student's t test |
| | mean | SD | n | mean | SD | n | *p* |
| | 942.0 | 99.5 | 3 | 921.2 | 97.8 | 5 | 0.7819 NS |
| Adult MPOA | *Sema6a*$^{+/+}$ | | | *Sema6a*$^{-/-}$ | | | Two-tailed unpaired Student's t test |
| | mean | SD | n | mean | SD | n | *p* |
| | 398.0 | 25.9 | 3 | 368.0 | 38.5 | 3 | 0.3258 NS |

**Supplementary Table 2.** Breeding records comparing *C57Bl/6J* to *Sema6a* knockout mice during 9 months of breeding.

| | *C57Bl/6J* ♂WT x ♀WT | *Sema6a* ♂HZ x ♀HZ | *Sema6a* ♂KO x ♀HZ/WT | **Sema6a* ♂KO x ♀HZ |
|---|---|---|---|---|
| Males used | 76 | 15 | 9 | 4 |
| Total matings | 290 | 86 | 20 | 8 |
| Litters | 233 | 59 | 7 | 4 |
| Males that produced a litter | 71 | 15 | 4 | 2 |
| Successful matings (% litters born of total matings) | 80.3 | 68.6 | 35.0 | 50.0 |
| Fertile males (% produced a litter of all males) | 93.4 | 100 | 44.4 | n.a. |
| Average litter size (number of pups) | 6.6 | 5.6 | 6.3 | 7.0 |

* This column represents subsequent breeding records only of the 4 fertile $Sema6a^{-/-}$ males.

**Supplementary Fig. 1**

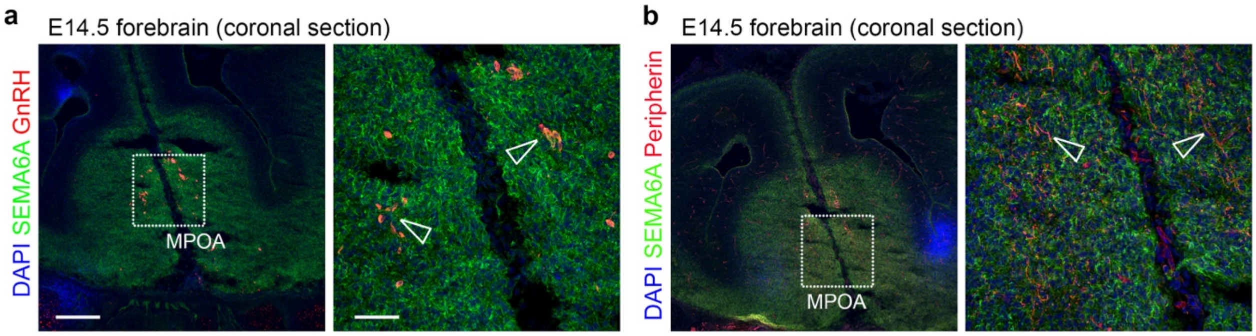

**Supplementary Fig. 1. SEMA6A is not expressed on GnRH neurons or TN axons in the forebrain.**

**(a)** Coronal sections of E14.5 mouse heads at the level of the MPOA were immunolabelled for SEMA6A (green) and GnRH (red). Empty arrowheads indicate lack of expression of SEMA6A in GnRH-positive neurons.

**(b)** Coronal sections of E14.5 mouse heads at MPOA level were immunolabelled for SEMA6A (green) and Peripherin (red). Empty arrowheads indicate lack of expression of SEMA6A on Peripherin-positive TN axons.

All sections were counterstained with DAPI. White dotted boxes indicate areas shown at higher magnification on the right of the corresponding panel.

Abbreviations: MPOA, medial preoptic area.

Scale bars: 200 µm (low magnification), 50 µm (high magnification).

### Supplementary Fig. 2

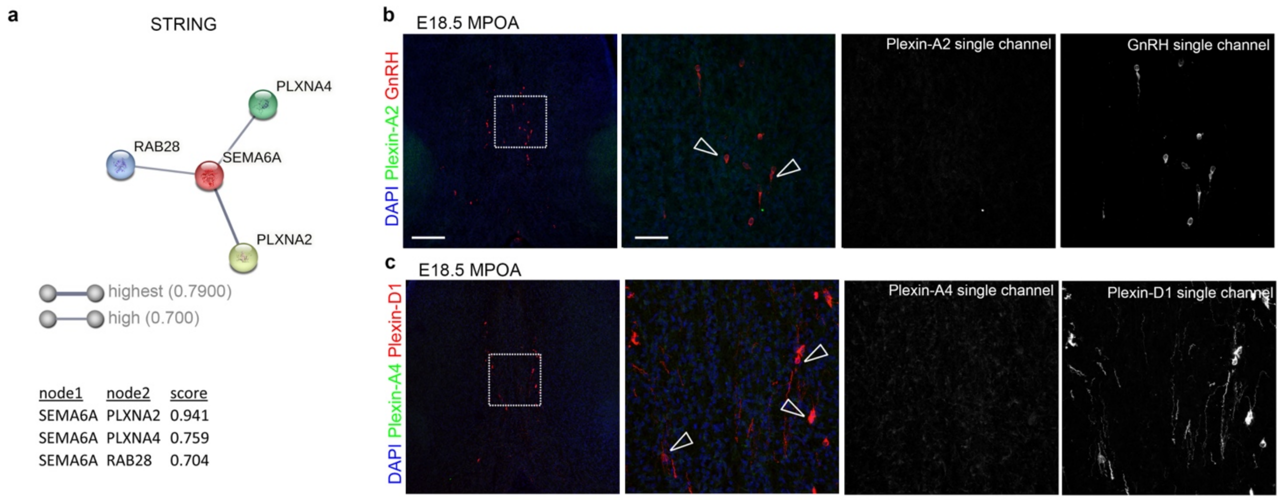

**Supplementary Fig. 2. Plexin-A2 and Plexin-A4 are not expressed on MPOA-resident GnRH neurons.**

**(a)** SEMA6A interactome computed using the STRING database of protein-protein interactions. A high confidence cut-off score of 0.700 was selected. The score for each protein pair is shown and the thickness of each network connection is proportional to the indicated interaction score value.

**(b)** Coronal sections of E18.5 mouse brains at the MPOA level were immunolabelled for Plexin-A2 (green) and GnRH (red). Single channels of magnified images are displayed on the right of each image. Empty arrowheads indicate lack of expression of Plexin-A2 on GnRH-positive neurons.

**(c)** Coronal sections of E18.5 mouse brains at the MPOA level were immunolabelled for Plexin-A4 (green) and Plexin-D1 (red). Single channels of magnified images are displayed on the right of each image. Empty arrowheads indicate lack of expression of Plexin-A4 on Plexin-D1-positive GnRH neurons.

All sections were counterstained with DAPI. White dotted boxes indicate areas shown at higher magnification on the right of the corresponding panel.

Abbreviations: MPOA, medial preoptic area.

Scale bars: 200 μm (low magnification), 50 μm (high magnification).

### Supplementary Fig. 3

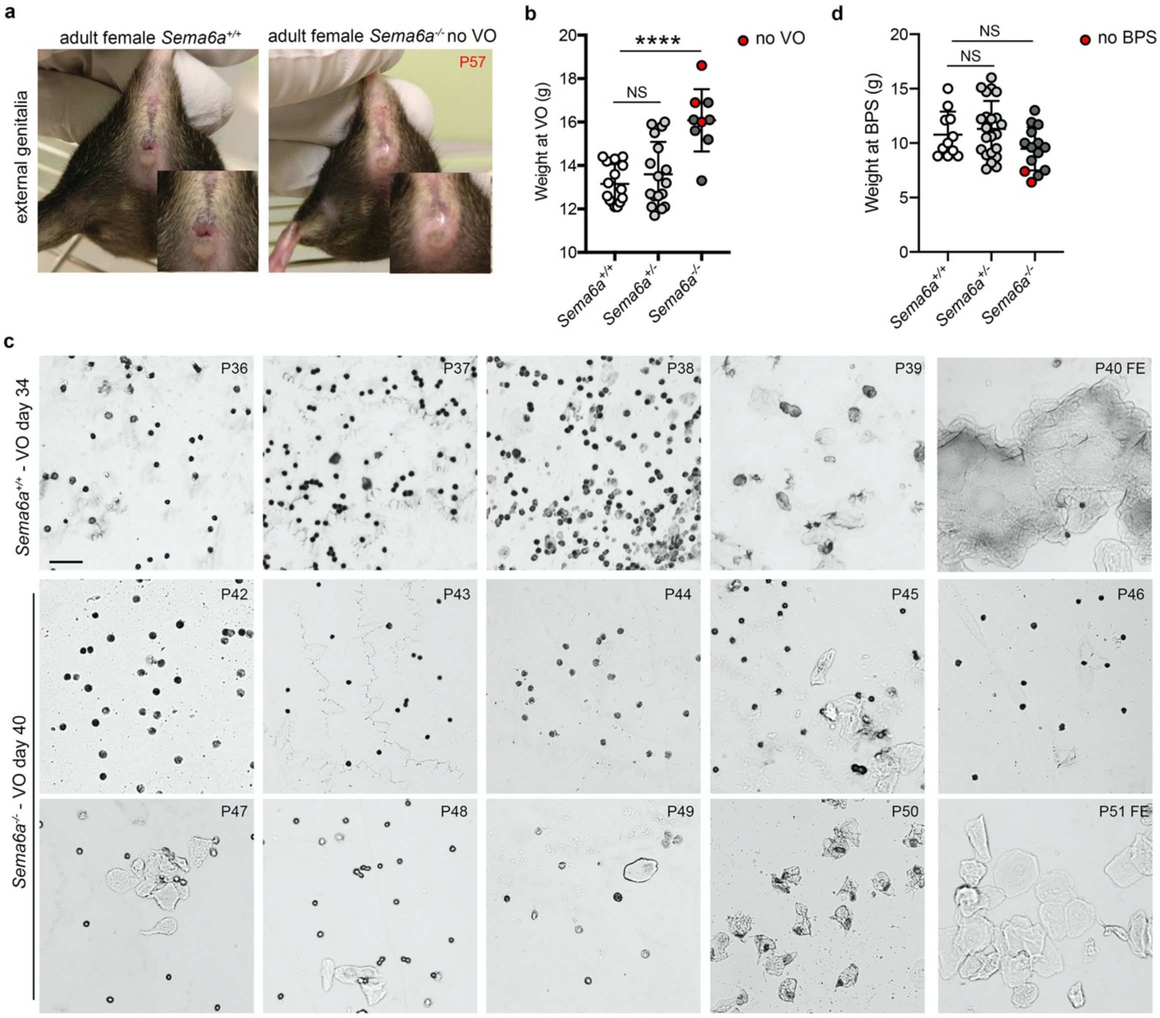

**Supplementary Fig. 3. Weight at puberty onset and first estrous assessment in *Sema6a*$^{-/-}$ mice.**

**(a)** Representative images of external genitalia of the indicated genotypes. *Sema6a*$^{+/+}$ female mice exhibit an opened vagina (left), whereas this *Sema6a*$^{-/-}$ female mouse did not reach VO at P57 (right). The inset at the bottom right shows higher magnification of the mouse vagina.

**(b)** Weight at the time of the vaginal opening (VO) in female mice of the indicated genotypes (*Sema6a*$^{+/+}$ n = 15; *Sema6a*$^{+/-}$ n = 17, $p = 0.5399$; *Sema6a*$^{-/-}$ n = 9, $p < 0.0001$). Red dots indicate *Sema6a*$^{-/-}$ female mice that did not show VO at the time the mouse was sacrificed.

**(c)** Representative images of vaginal smears of the indicated genotypes from 2 days after VO to the appearance of first estrous (FE). FE was identified by presence of a majority of anucleated cornified epithelial cells.

**(d)** Weight at the time of balanopreputial separation (BPS) in adult male mice of the indicated genotypes (*Sema6a*$^{+/+}$ n = 11; *Sema6a*$^{+/-}$ n = 22, $p = 0.7399$; *Sema6a*$^{-/-}$ n = 14, $p = 0.2725$). Red dots indicate *Sema6a*$^{-/-}$ male mice that did not show BPS by the time of sacrifice.

**** $p < 0.0001$ and NS, not significant after One-way ANOVA followed by Dunnett's post-hoc test.

Abbreviations: VO, vaginal opening; BPS, balanopreputial separation.

Scale bar: 100 μm.

Data are presented as mean ± SD. Source data are provided as a Source Data file

**Supplementary Fig. 4**

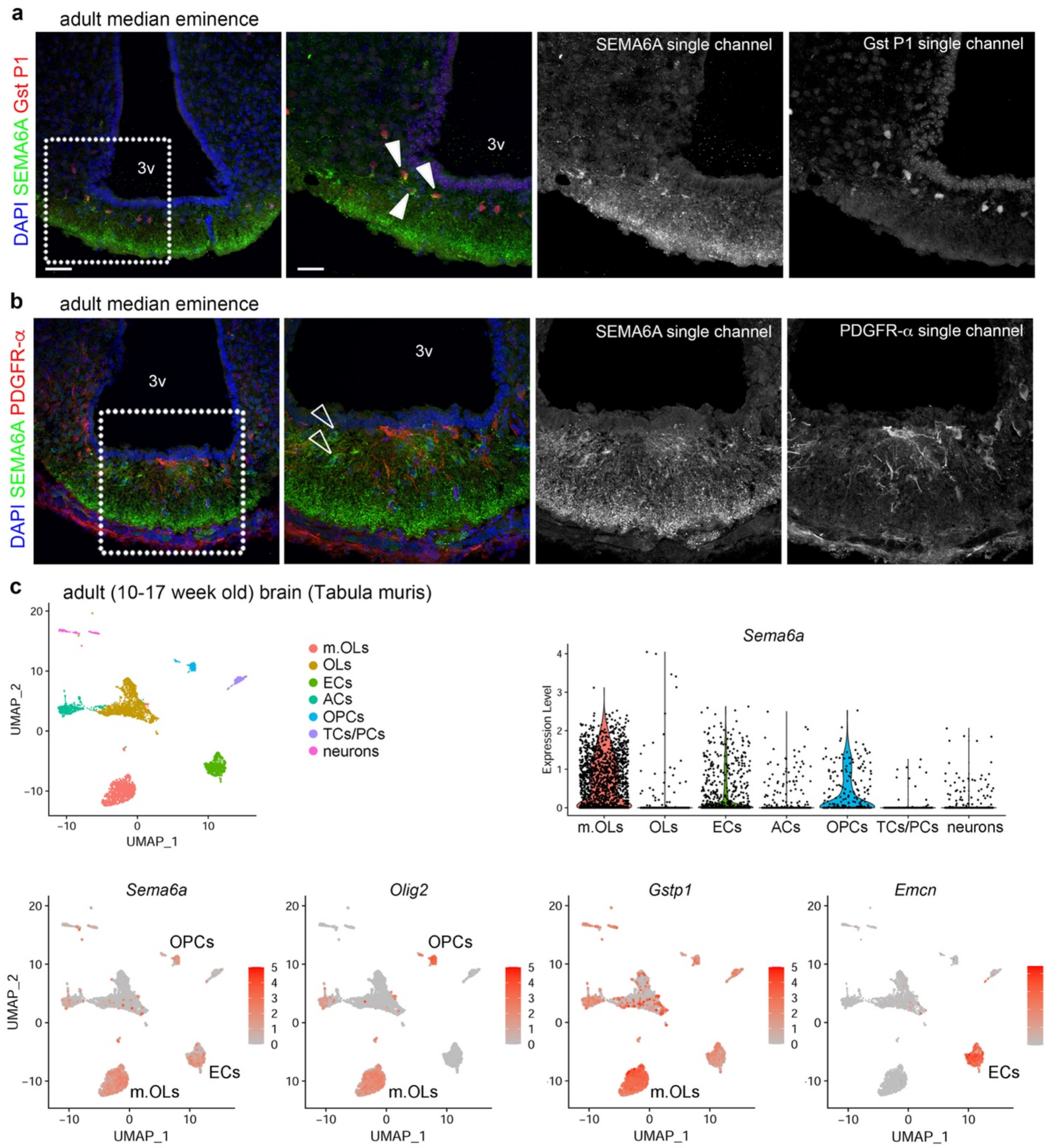

**Supplementary Fig. 4. SEMA6A is expressed by maturing oligodendrocytes (OLs).**

**(a)** Coronal sections of adult mouse brains showing the ME immunolabelled for SEMA6A (green) and Gst P1 (red) to label maturing OLs. Single channels of images at higher magnification are displayed on the right of each image. Solid arrowheads indicate the expression of SEMA6A on Gst P1$^{+}$ maturing OLs.

**(b)** Coronal sections of adult mouse brains showing the ME immunolabelled for SEMA6A (green) and PDGFR-α (red) to label OPCs. Single channels of images at higher magnification are displayed on the right of each image. Empty arrowheads indicate the lack of expression of SEMA6A on PDGFR-α$^+$ OPCs.

**(c)** scRNA-seq analysis of the adult mouse brain from the Tabula Muris dataset. UMAP plots show distinct cell types (top left panel) and *Sema6a, Olig2*, *Gstp1* and *Emcn* transcript levels (bottom panels). Violin plots compare *Sema6a* transcript levels (top right panel) in the different brain cell populations.

All sections were counterstained with DAPI. White dotted boxes indicate areas shown at higher magnification on the right of the corresponding panel.

Abbreviations: 3v, third ventricle; m.OLs, maturing OLs; OPCs, oligodendrocyte precursor cells; ECs, endothelial cells; ACs, astrocytes; TCs/PCs, tanycytes/pericytes.

Scale bars: 50 μm (low magnification), 25 μm (high magnification).

### Supplementary Fig. 5

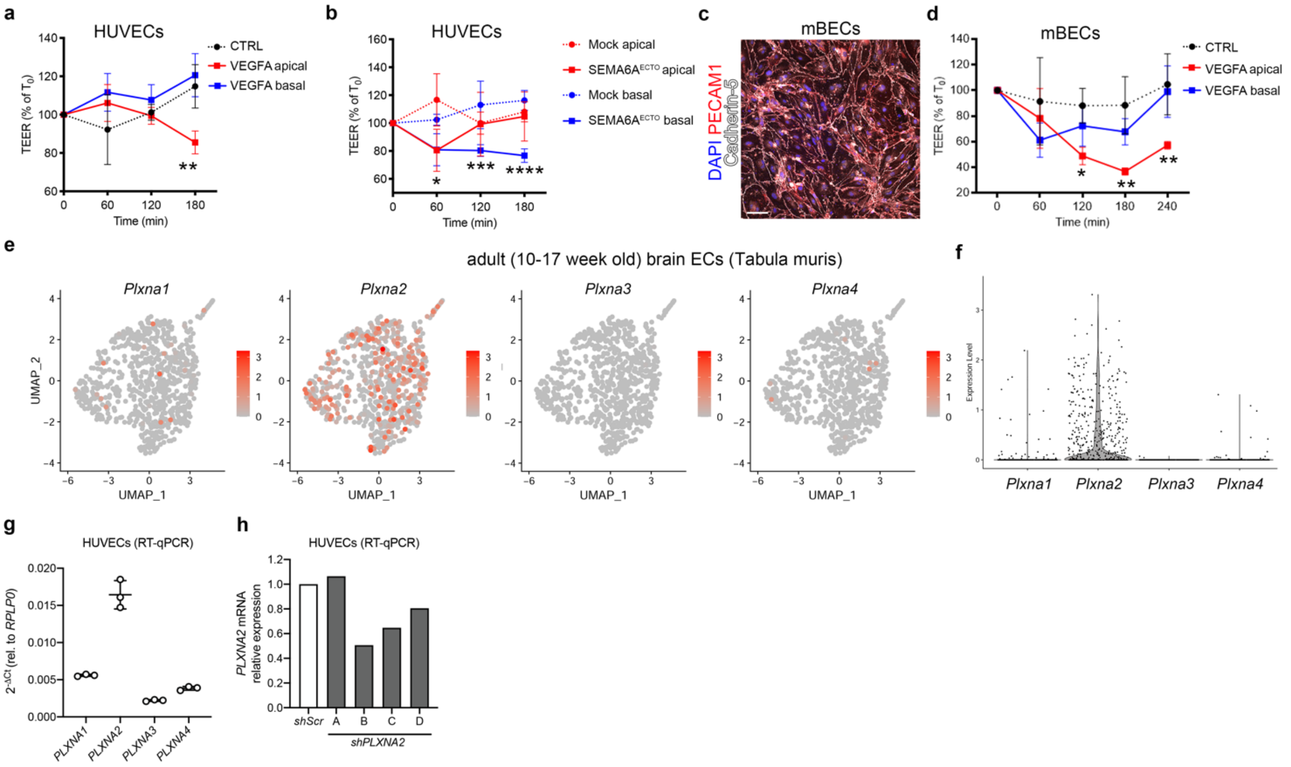

**Supplementary Fig. 5. Abluminal SEMA6A regulates vascular permeability via Plexin-A2 in human and mouse ECs.**

**(a)** TEER quantification in HUVECs apically (red) or basally (blue) treated with control (CTRL) or VEGF-A-containing media (50 ng/mL). TEER values are expressed as percentage of TEER at time 0 ($T_0$), ranging 10-15 Ωcm$^2$ (VEGF-A apical vs CTRL: 60 min $p$ = 0.1474, 120 min $p$ = 0.9647, 180 min $p$ = 0.0012; VEGFA basal vs CTRL: 60 min $p$ = 0.0304, 120 min $p$ = 0.6442, 180 min $p$ = 0.6929). Graph shows one out of n = 3 independent experiments (n = 3 per group).

**(b)** TEER quantification in HUVECs apically (red) or basally (blue) treated with conditioned media from SEMA6A$^{ECTO}$ or mock-transfected COS-7 cells. TEER values are expressed as percentage of TEER at time 0 ($T_0$), ranging 7-21 Ωcm$^2$. Graph shows one out of n = 3 independent experiments (at least n = 3 per group).

**(c)** mBECs were immunolabeled for PECAM1 (red), Cadherin-5 (white) to assess purity of primary cultured cells. Nuclei were counterstained with DAPI.

**(d)** TEER quantification in mBECs apically (red) or basally (blue) treated with control (CTRL) or VEGFA-containing media (50 ng/mL). TEER values are expressed as percentage of TEER at time 0 ($T_0$), ranging 42-67 Ωcm$^2$ (VEGFA apical vs CTRL: 60 min $p$ = 0.5842, 120 min $p$ = 0.0157, 180 min $p$ = 0.0014, 240 min $p$ = 0.0033; VEGFA basal vs CTRL: 60 min $p$

= 0.0740, 120 min *p* = 0.4705, 180 min *p* = 0.2781, 240 min *p* = 0.9040). Graph shows one out of n = 2 independent experiments (at least n = 3 per group).

**(e,f)** scRNA-seq analysis of adult mouse brain ECs from the Tabula Muris dataset. UMAP plots (**e**) and violin plots (**f**) show *Plxna1*, *Plxna2*, *Plxna3* and *Plxna4* transcript levels in the EC subset.

**(g)** RT-qPCR analysis for *PLXNA1-4* transcripts in HUVECs. mRNA levels were calculated relative to control scramble shRNA (*shScr*) samples using *RPLP0*-normalized Ct threshold values.

**(h)** RT-qPCR analysis for *PLXNA2* transcript in HUVECs after lentiviral infection with *shScr* and indicated *PLXNA2* shRNA (*shPLXNA2* A-D) (n = 1 per group). *PLXNA2* mRNA levels were calculated relative to control *shScr* samples using *RPLP0*-normalized Ct threshold values.

* $p < 0.05$, ** $p < 0.01$, *** $p < 0.001$, **** $p < 0.0001$ after Two-way ANOVA followed by Tukey post-hoc test.

Abbreviations: ECs, endothelial cells; mBECs, mouse brain endothelial cells; TPM, transcript per million.

Scale bar: 100 µm (c).

Data are presented as mean ± SD. Source data are provided as a Source Data file**.**

**Supplementary Fig. 6**

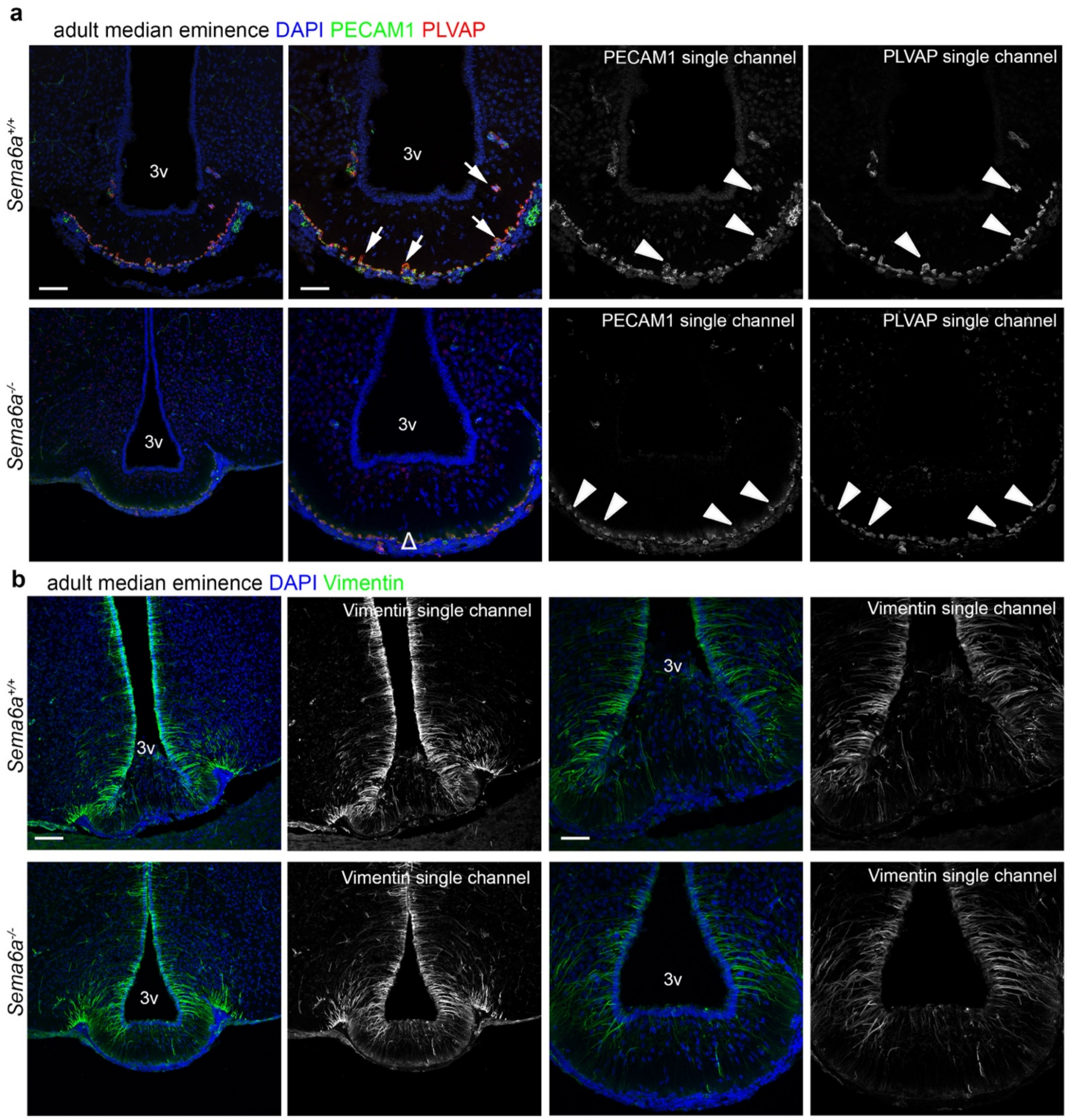

**Supplementary Fig. 6. Loss of SEMA6A induces general vascular remodeling.**

**(a)** Coronal sections of adult female brains with the indicated genotypes at the level of the ME were immunolabelled for PECAM1 (green) and PLVAP (red) to label ME blood vessels and fenestrated capillaries, respectively. Single channels of images at higher magnification are displayed on the right of each image. White arrows indicate normal presence of PECAM1$^+$/PLVAP$^+$ capillary loops in *Sema6a*$^{+/+}$ mice which are lacking in *Sema6a*$^{-/-}$ (Δ) mice. Solid arrowheads indicate that ME blood vessel in both genotype express pan-endothelial marker PECAM1 and capillary fenestrae marker PLVAP.

**(b)** Coronal sections of adult female brains with the indicated genotypes at the level of the ME were immunolabelled for Vimentin (green) to label β2-tanycytes. Single channels of images at higher magnification are displayed on the right of each image.
All sections were counterstained with DAPI. White dotted boxes indicate areas shown at higher magnification on the right of the corresponding panel.
Abbreviations: 3v, third ventricle.
Scale bars: 100 µm (low magnification), 50 µm (high magnification).

**Supplementary Fig. 7**

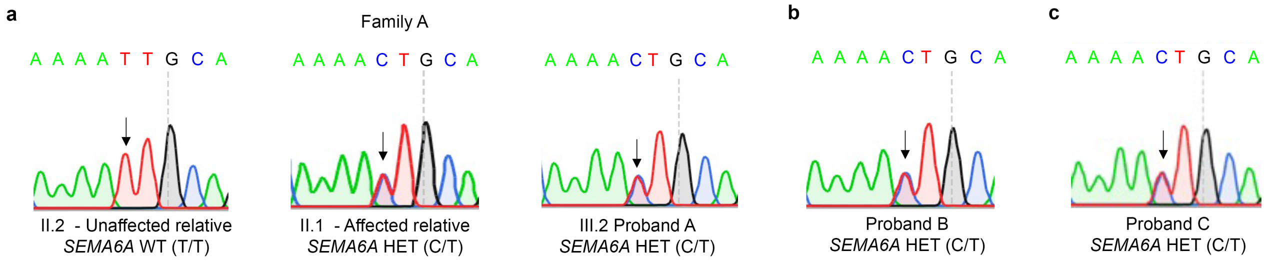

**Supplementary Fig. 7. Sanger sequencing of probands confirms the presence of the *SEMA6A* heterozygous variant.**

**(a)** Electropherograms of index family A members. Proband A (III.2, right) and the affected relative (II.1, center) showed the c.1268T>C substitution in heterozygous state (T/C), while the unaffected relative (II.2, left) is homozygous for the reference nucleotide (T/T).

**(b, c)** Electropherograms of Proband B (b) and Proband C (c). Both probands showed the c.1268>C substitution in heterozygous state (T/C).

Abbreviations: WT, wild type; HET, heterozygous.
